# Evaluating the Concordance between International Classification of Diseases, Tenth Revision Code and Stroke Severity as Measured by the National Institutes of Health Stroke Scale

**DOI:** 10.1101/2024.02.21.24303177

**Authors:** Mohamed Taha, Mamoon Habib, Victor Lomachinsky Torres, Peter Hadar, Joseph P Newhouse, Lee H. Schwamm, Deborah Blacker, Lidia MVR Moura

**Affiliations:** Department of Neurology, Massachusetts General Hospital, Harvard Medical School, Boston, Massachusetts; Department of Health Care Policy, Harvard Medical School, Boston, Massachusetts. Mongan Institute, Massachusetts General Hospital, Boston, Massachusetts; National Bureau of Economic Research, Cambridge, Massachusetts; Department of Health Policy and Management, Harvard T.H. Chan School of Public Health, Boston, Massachusetts; Harvard Kennedy School, Cambridge, Massachusetts; Digital Strategy and Transformation, Office of the Dean, Yale School of Medicine; Biomedical Informatics & Data Sciences at Yale School of Medicine; Department of Epidemiology, Harvard T.H. Chan School of Public Health, Boston, Massachusetts; Department of Psychiatry, Massachusetts General Hospital, Boston, Massachusetts; Department of Psychiatry, Harvard Medical School, Boston, Massachusetts

**Keywords:** National Institutes of Health Stroke Scale, NIHSS, International Classification of Diseases Tenth Revision Code, ICD-10, concordance between NIHSS and ICD10, Paul Coverdell National Acute Stroke Program, Medicare claims data

## Abstract

**Background:** The National Institutes of Health Stroke Scale (NIHSS) scores have been used to evaluate Acute Ischemic Stroke (AIS) severity in clinical settings. Through the International Classification of Diseases, Tenth Revision Code (ICD-10), documentation of NIHSS scores has been made possible for administrative purposes and has since been increasingly adopted in insurance claims. Per CMS guidelines, the stroke ICD-10 diagnosis code must be documented by the treating physician, but ICD-10 NIHSS scores can be documented by any healthcare provider involved in the patient’s care. Accuracy of the administratively collected NIHSS compared to expert clinical evaluation as documented in the Paul Coverdell registry is however still uncertain.

**Methods:** Leveraging a linked dataset comprised of the Paul Coverdell National Acute Stroke Program (PCNASP) clinical registry and probabilistically matched individuals on Medicare Claims data, we sampled patients aged 65 and above admitted for AIS across nine states, from 2016 to 2019. We excluded those lacking documentation for either clinical or ICD-10 based NIHSS scores. We then examined score concordance from both databases and measured discordance as the absolute difference between the PCNASP and ICD-10-based NIHSS scores.

**Results:** Among 66,837 matched patients, mean NIHSS scores for PCNASP and Medicare ICD-10 were 7.26 (95% CI: 7.20 – 7.32) and 7.40 (95% CI: 7.34 – 7.46), respectively. Concordance between the two scores was high as indicated by an intraclass correlation coefficient of 0.93.

**Conclusion:** The high concordance between clinical and ICD-10 NIHSS scores highlights the latter’s potential as measure of stroke severity derived from structured claims data.

## Introduction

The National Institutes of Health Stroke Scale (NIHSS) has long served as a vital tool for assessing the severity of acute ischemic strokes^1^. This well-established instrument has been employed in numerous clinical trials related to stroke and has garnered validation as a reliable predictor of stroke outcomes^2–4^. Beyond its direct clinical utility, hospitals have also been recording the NIHSS score for administrative risk adjustment and claims purposes, which was enabled by the tenth revision of the International Classification of Diseases (ICD) codes^5^.

However, during the initial national optional reporting period in the United States, NIHSS scores were documented through administrative claims in only 15% of hospital admissions for acute ischemic stroke^5^.

A notable gap in the existing literature pertains to an evaluation of the accuracy of ICD-based NIHSS scores when compared to clinical NIHSS scores. The incorporation of ICD-10 based NIHSS is advocated for its effectiveness in hospital risk adjustment^6^ and in enhancing the measurement of quality of care and patient outcomes^7^. A recent study took a step in this direction by assessing the validity of ICD-10 based NIHSS scores using data from a single center stroke registry^8^. While this study revealed a high level of agreement between ICD-10 based NIHSS scores and clinical NIHSS scores (R2 = 0.86), it is worth noting that ICD-10 NIHSS scores were available for only a limited subset of patients (n =395 out of 1357, 29.1%).

Furthermore, it is essential to recognize that single institution studies do not fully capture the potential diversity of ICD coding practices across different states and patient populations ^8^. In light of these considerations, we examined the concordance between the NIHSS score reported for clinical assessment in the Paul Coverdell National Acute Stroke Program (PCNASP) and the score reported in Medicare claims database, with data gathered over multiple years across 9 states.

### Objective

To evaluate the concordance between Acute Ischemic Stroke (AIS) severity scores from the clinically derived NIHSS scores obtained from PCNASP registry and administratively derived ICD-10 NIHSS scores obtained from Medicare claims.

## Methodology

### Data Source

We established a linkage between the Paul Coverdell National Acute Stroke Program (PCNASP) registry and Medicare Claims data. PCNASP aims to collect and track data on stroke-related cases to enhance patient care quality^9^. The PCNASP registry includes data from 2008 to 2020 across 9 US states (California, Georgia, Massachusetts, Michigan, Minnesota, New York, Ohio, Washington, and Wisconsin), and captures the NIHSS scores as reported by clinicians or hospital staff.

Medicare, a national health insurance program administered by the Centers for Medicare & Medicaid Services (CMS), primarily serves individuals aged 65 or older. From October 2016, Medicare implemented the use of ICD10’s R29.7xx codes to group NIHSS scores creating distinct severity categories that aid stroke outcome research^10^. The Medicare Provider Analysis and Review (MEDPAR) database contains extensive information about beneficiaries and as well a broad range of information, including patient demographics, admission and discharge dates, diagnosis, and procedure codes, provider identifiers, comorbidities, and, since 2016, measurement of stroke severity through ICD-10 based NIHSS scores^11^. This dataset is utilized for administrative purposes s well as for healthcare research.

### Linking Databases

Due to the lack of common unique patient identifiers across the two databases, we applied a validated probabilistic matching strategy to link individuals in the PCNASP and Medicare datasets^12^. This approach utilized variables such as age, gender, admission and discharge dates, diagnosis code, hospitals, and state. After the linking process, we retained only those patients with unique matches, excluding cases where PCNASP IDs corresponded to multiple Medicare Beneficiary IDs, and vice versa.

### Study Population

With the linked dataset, we selected patients aged 65 or older who were hospitalized for AIS and discharged under ICD-10 codes within the I63 groups. ^13^. Patients were required to have both PCNASP (clinical) and Medicare (ICD-10-based) NIHSS scores recorded. Since the ICD-10-based NIHSS score codes were introduced with the ICD-10th revision, we limited our analysis to cases from 2016 to 2019 that used ICD-10 diagnosis and procedure codes. The cut-off in 2019 was chosen to avoid the COVID19 era and beyond

### Measurements

NIHSS scores are the primary outcome measured. The scores range from 0-42 with increasing values denoting more severe deficits^14^. In ICD-10, the R29.7xx code group allows for direct input of integer values in that range and reflects the identical scoring criteria.

Per CMS guidelines, while stroke ICD-10 diagnosis code must be documented by the treating physician, documentation of ICD-10 NIHSS scores may be based on medical record documentation from any healthcare providers involved in the patient’s care^15^.

### Analysis Plan

We assessed the concordance between the NIHSS scores documented in PCNASP and in Medicare claims data in the MEDPAR files. For our analysis in Medicare data, our main emphasis was on the ICD-10 NIHSS score documented at the moment of admission. Any instances where multiple ICD-10 NIHSS scores were present at admission or where there was no ICD-10 NIHSS score at admission were excluded from our analysis.

### Statistical Analysis

Our analysis focused on evaluating the concordance between the ordinal values of the PCNASP and Medicare NIHSS scores. We assessed the concordance of these two scores using the Intraclass Correlation Coefficient (ICC3) with a two-way mixed-effects model^16^. Lastly, we calculated discordance as the absolute difference between the the PCNASP and Medicare NIHSS scores. Our study adhered to the ethical guidelines outlined by the Massachusetts General Hospital Institutional Review Board (IRB) and obtained IRB approval under the protocol number 2020P003963. We followed the Strengthening the Reporting of Observational Studies in Epidemiology (STROBE) guidelines for observational studies^17^ (Supplementary materials).

## Results

From an initial dataset of 233,908 patients, 66,837 patients with 67,197 AIS admissions met our inclusion criteria. These patient data came from 477 hospitals across nine states, with state-specific demographics and results provided in the Supplementary materials (Table S1)

Table 1 details the breakdown of PCNASP (clinical) and Medicare (ICD-10-based) NIHSS scores and their demographic characteristics. The mean age was 79.1 ± 8.67 years. Our population was predominantly white (81.0%).

**Table 1:**
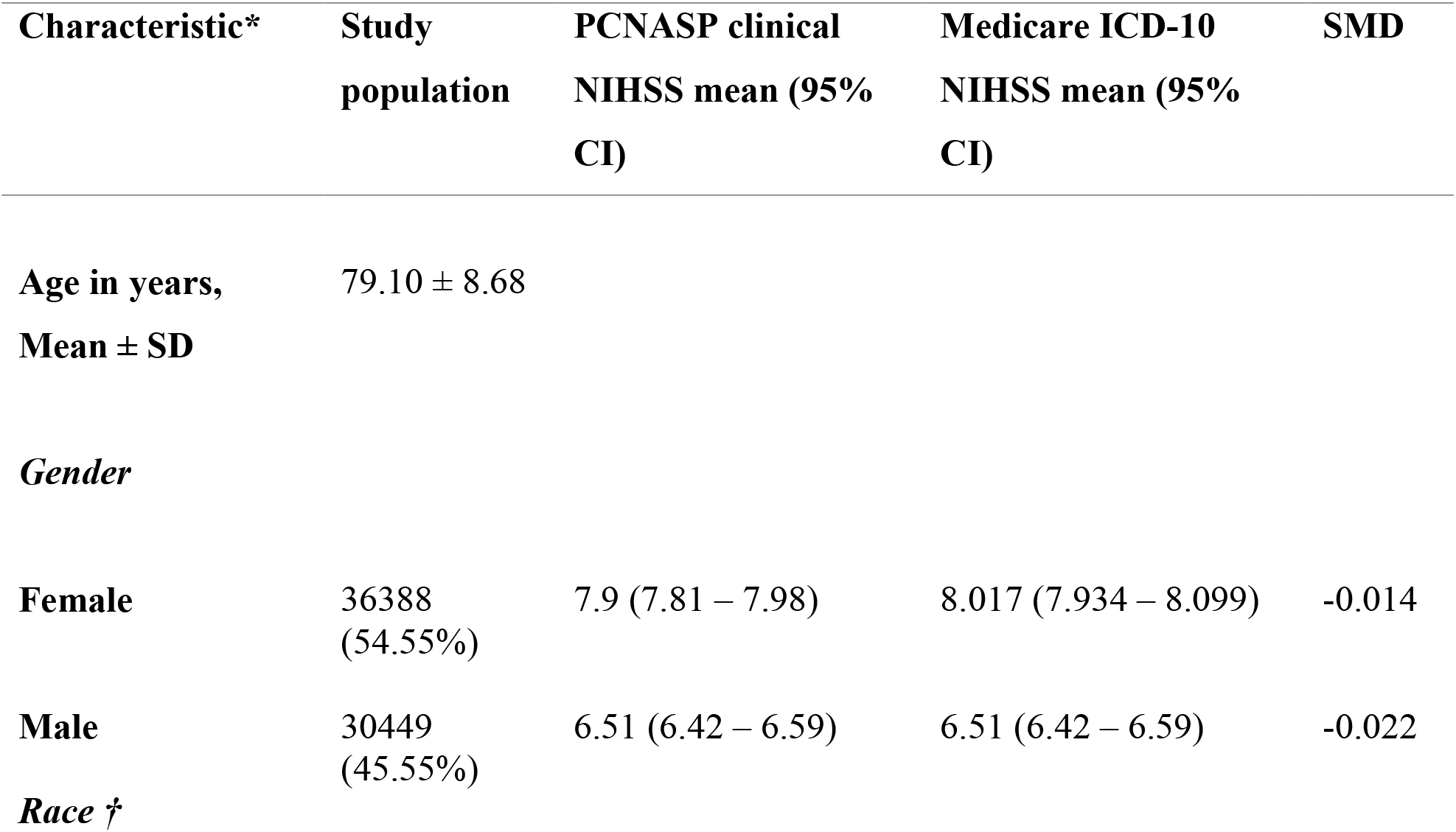
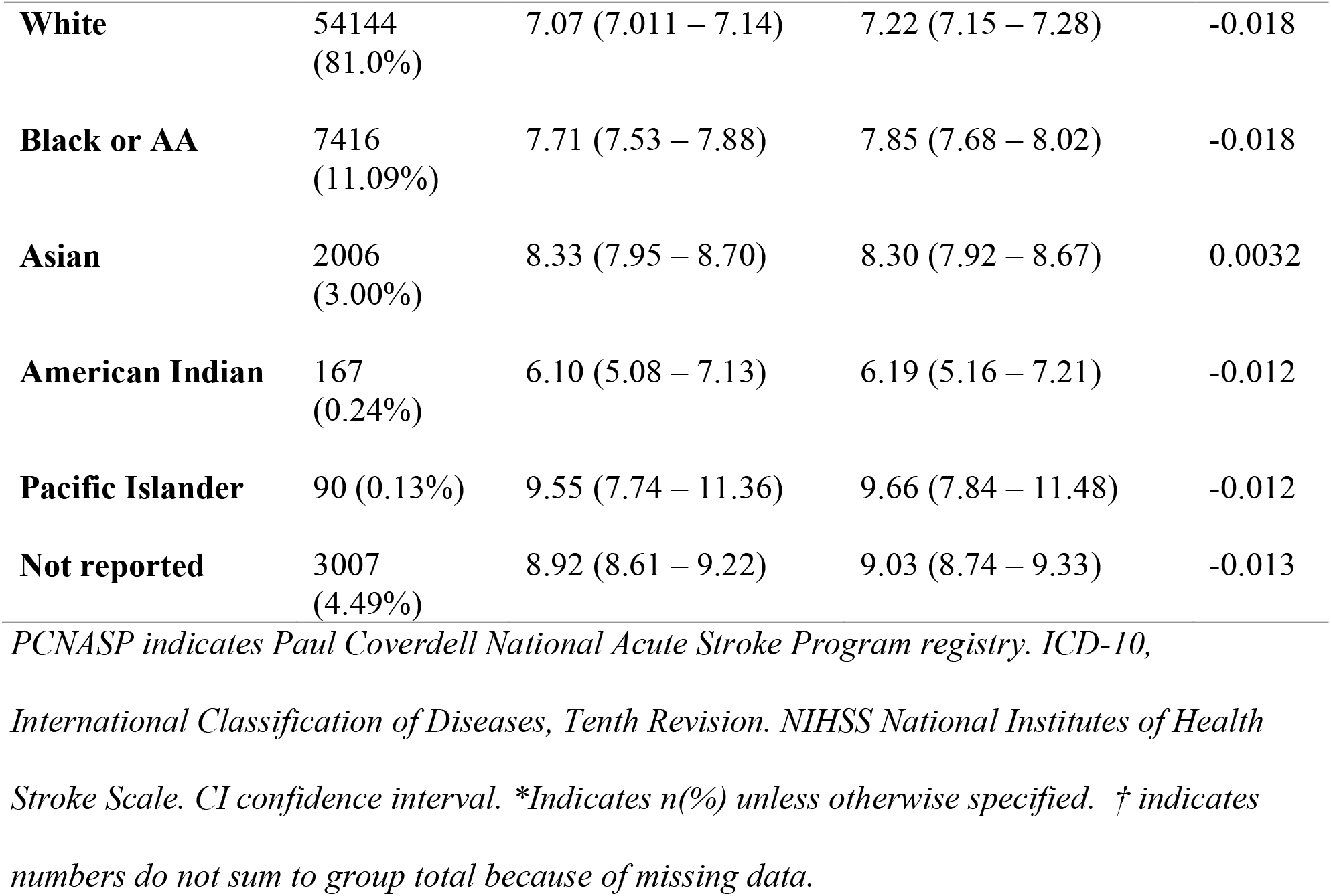
Study cohort demographics and average clinical and ICD-10 NIHSS scores.

| Characteristic* | Study population | PCNASP clinical NIHSS mean (95% CI) | Medicare ICD-10 NIHSS mean (95% CI) | SMD |
| --- | --- | --- | --- | --- |
| **Age in years, Mean ± SD** | 79.10 ± 8.68 | | | |
| **Gender** |  |  |  |  |
| **Female** | 36388 (54.55%) | 7.9 (7.81 – 7.98) | 8.017 (7.934 – 8.099) | -0.014 |
| **Male** | 30449 (45.55%) | 6.51 (6.42 – 6.59) | 6.51 (6.42 – 6.59) | -0.022 |
| **Race †** |  |  |  |  |
| **White** | 54144<br>(81.0%) | 7.07 (7.011 – 7.14) | 7.22 (7.15 – 7.28) | -0.018 |
| **Black or AA** | 7416<br>(11.09%) | 7.71 (7.53 – 7.88) | 7.85 (7.68 – 8.02) | -0.018 |
| **Asian** | 2006<br>(3.00%) | 8.33 (7.95 – 8.70) | 8.30 (7.92 – 8.67) | 0.0032 |
| **American Indian** | 167<br>(0.24%) | 6.10 (5.08 – 7.13) | 6.19 (5.16 – 7.21) | -0.012 |
| **Pacific Islander** | 90 (0.13%) | 9.55 (7.74 – 11.36) | 9.66 (7.84 – 11.48) | -0.012 |
| **Not reported** | 3007<br>(4.49%) | 8.92 (8.61 – 9.22) | 9.03 (8.74 – 9.33) | -0.013 |

*PCNASP indicates Paul Coverdell National Acute Stroke Program registry. ICD-10,*
*International Classification of Diseases, Tenth Revision. NIHSS National Institutes of Health*
*Stroke Scale. CI confidence interval. \*Indicates n(%) unless otherwise specified. † indicates*
*numbers do not sum to group total because of missing data.*

The mean PCNASP clinical NIHSS score was 7.26 (95%CI: 7.20 – 7.32), while the mean Medicare ICD-10-based NIHSS score was 7.40 (95%CI: 7.34 – 7.46). Both scores had the same median of 4.0 with interquartile range of (2.0 - 11.0).

The two scores demonstrated high concordance, as indicated by ICC3 of 0.93. As illustrated in Figure 1, the Bland-Altman plot demonstrates a minimal mean difference between the clinical and ICD-10 NIHSS scores, consistent with a high level of agreement rates.

**Figure 1:**
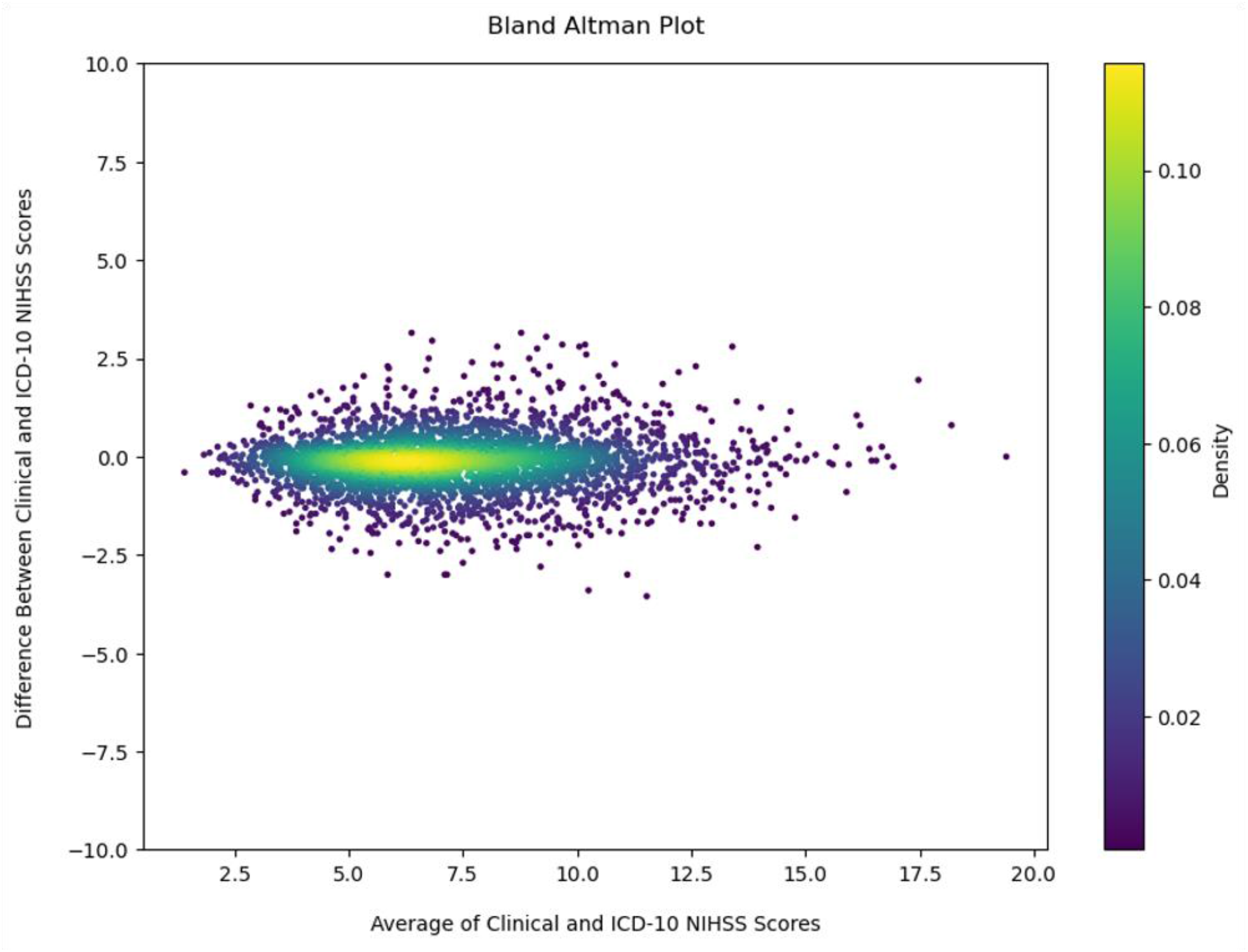
This Bland-Altman plot illustrates the concordance between the clinical and ICD-10 NIHSS scores. The y-axis represents the difference between the two scores, while the x-axis shows the average score obtained by combining them and dividing by 2. Data points, each representing an aggregate of 20 individual cases to enhance visual interpretability, form a heat map indicating the density of sample differences. The color gradient from yellow to purple visually encodes the sample density, providing an immediate sense of the distribution of agreement across the score range. There is a total of 3252 data points shown. The central band of the plot, highlighted in yellow-green, denotes the area where the majority of the sample scores are tightly clustered within ±1.96 standard deviations of the mean difference—indicating a strong agreement between the two scales. Areas of data points in purple, dispersed further from the mean, signify fewer occurrences, representing a greater discrepancy in scores.

**Figure 2:**
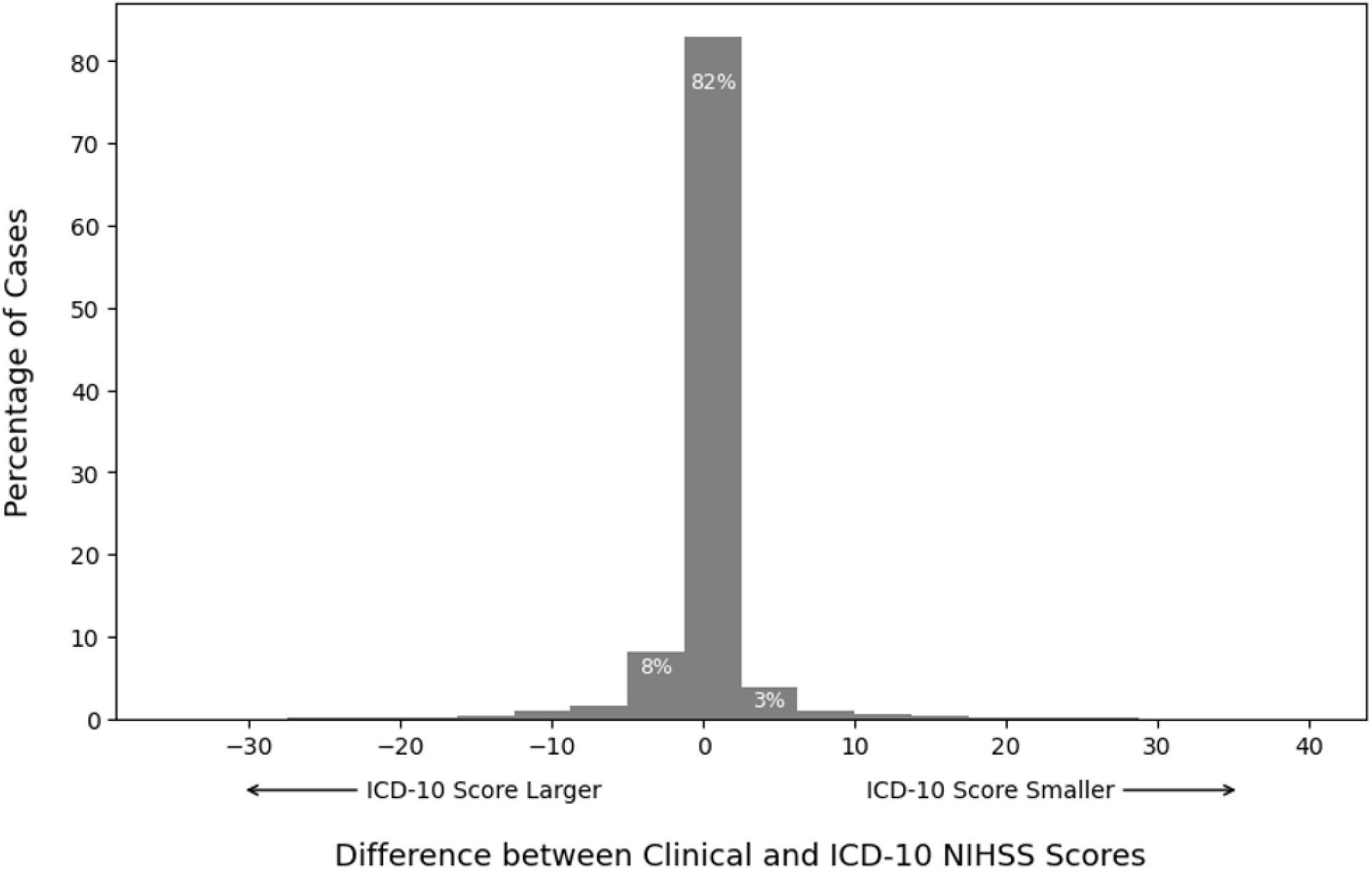
Discrepancy Between Clinical and ICD-10 NIHSS Scores. This figure shows the percentage of cases with differences between clinical NIHSS scores and ICD-10 codes. The central gray bar shows that in 82% of cases the scores match. When disagreement occurs between the two scores, ICD-10 tends to be larger than clinical NIHSS.

In state-specific data, New York led in AIS cases (n = 15,080), followed by Ohio (n = 11,364), and Washington with the lowest count (n = 3,568) (Table S1). Notably, California recorded the highest mean clinical and ICD-10 NIHSS scores at 8.32 and 8.29, respectively, while Minnesota reported the lowest clinical NIHSS of 6.44, and Massachusetts the lowest ICD-10 NIHSS of 6.52.

The discordance, which is the absolute difference between the PCNASP and Medicare NIHSS scores, was 1.10 points (95% CI: 1.08 – 1.12) for the cohort. This discordance varied by state, with the highest average absolute score difference observed in California with 1.55 points (95% CI: 1.47 – 1.63) and the lowest in Massachusetts with 0.61 points (95% CI: 0.55 – 0.65) (Table S1).

## Discussion

In our investigation involving 66,837 AIS patients from 477 hospitals across nine states, we probed the concordance between clinical NIHSS score as registered in the Paul Coverdell Registry and its ICD-10-based administrative records counterpart in Medicare claims. Despite state-specific variations, our cohort consistently demonstrated an overall high concordance score of 0.93. Notably, our findings aligned with a smaller study that highlighted minimal discordance between the two scores in the state of New York^8^.

Bland-Altman plot revealed that the association between stroke severity and documentation concordance was non-linear. Particularly, we observed greater consistency in documentation at the extreme ends of the stroke severity spectrum. This could be due to the clinical characteristics and consequences of the stroke being more distinct and easier to document accurately at the extreme ends of the severity spectrum.

Our study identified regional and state-specific discordance discrepancies, partly explained by variations in patient populations and demographics. However, it is worth stressing that participation in the PCNASP database is voluntary and different numbers of hospitals participated in each state over time. Additionally, potential differences in stroke assessment, and in the ways through which claims are filed to Medicare by providers could also vary by location and practice ^18–20^.

While our approach of treating stroke severity score as a ordinal variable facilitated a rigorous statistical analysis, it’s essential to acknowledge that the slight variations we observed may not carry substantial clinical significance. Nevertheless, these documentation differences could become significant in borderline values where minimal score variations would result in change in stroke severity with subsequent reimbursement repercussions.

In addition, it is likely that with higher clinical NIHSS scores, larger absolute numerical variances can be noted as there is more granularity (i.e., more points) available in the score for strokes of greater. Therefore, larger variations for patients with higher scores may carry the same clinical impact as smaller variations in the less severe cases. **Limitations**

Our study predominantly analyzed data from a substantial population sample spanning 2016 to 2019, a period preceding the COVID-19 pandemic. This timeframe allows for an insightful examination of stroke-related documentation and claims in a pre-pandemic context. It is important to note, however, that stroke presentations, outcomes, and associated documentation practices have evolved during and post-COVID^21–24^. While these changes may influence the applicability of our findings, they remain relevant for understanding pre-pandemic trends and could offer valuable insights for future healthcare practices.

Moreover, our outcome measurements within the pre-pandemic period are not without limitations. The reliance on administrative documentation, often constrained by time and system complexities, introduces potential inaccuracies^10,18,25–27^. These limitations underscore the need for cautious interpretation of our findings. Furthermore, the generalizability of our results is restricted to the healthcare settings and regions included in our study. Variations in patient demographics, stroke care protocols, and documentation practices across different healthcare institutions and regions may limit the applicability of our findings to other contexts.

## Conclusion

Our study revealed a remarkable alignment between clinically assessed NIHSS scores and ICD-10 NIHSS scores derived through administrative means, thus bolstering the validity of the latter as a viable proxy for gauging stroke severity. Although certain clinical and demographic factors did exhibit a slight discordance, their practical clinical significance is arguably minimal.

Conducting extensive, multi-center analyses on a larger scale with the ICD-10 NIHSS score serving as a gauge of stroke severity can provide valuable insights into stroke epidemiology and further advancements in stroke management.

## Data Availability

Data are not available for public use

List: Non-standard Abbreviations and Acronyms

## Abbreviation Meaning

AIS: Acute Ischemic Stroke
ICD-10: International Classification of Diseases, Tenth Revision Code
LOS: Length of Hospital Stay
NIHSS: National Institutes of Health Stroke Scale
PCNASP: Paul Coverdell National Acute Stroke Program

## Tables and Figures

STROBE Statement—checklist of items that should be included in reports of observational studies

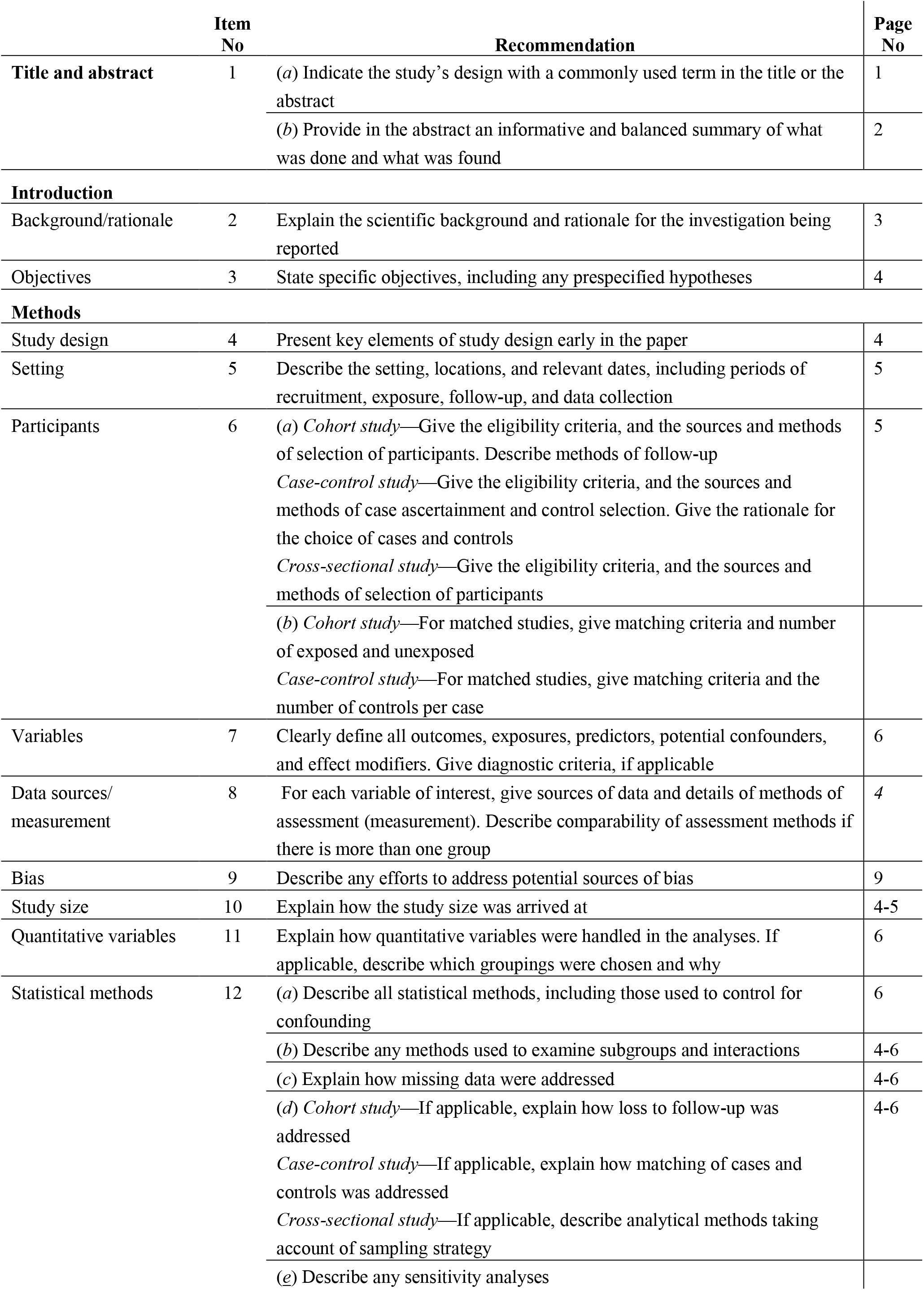
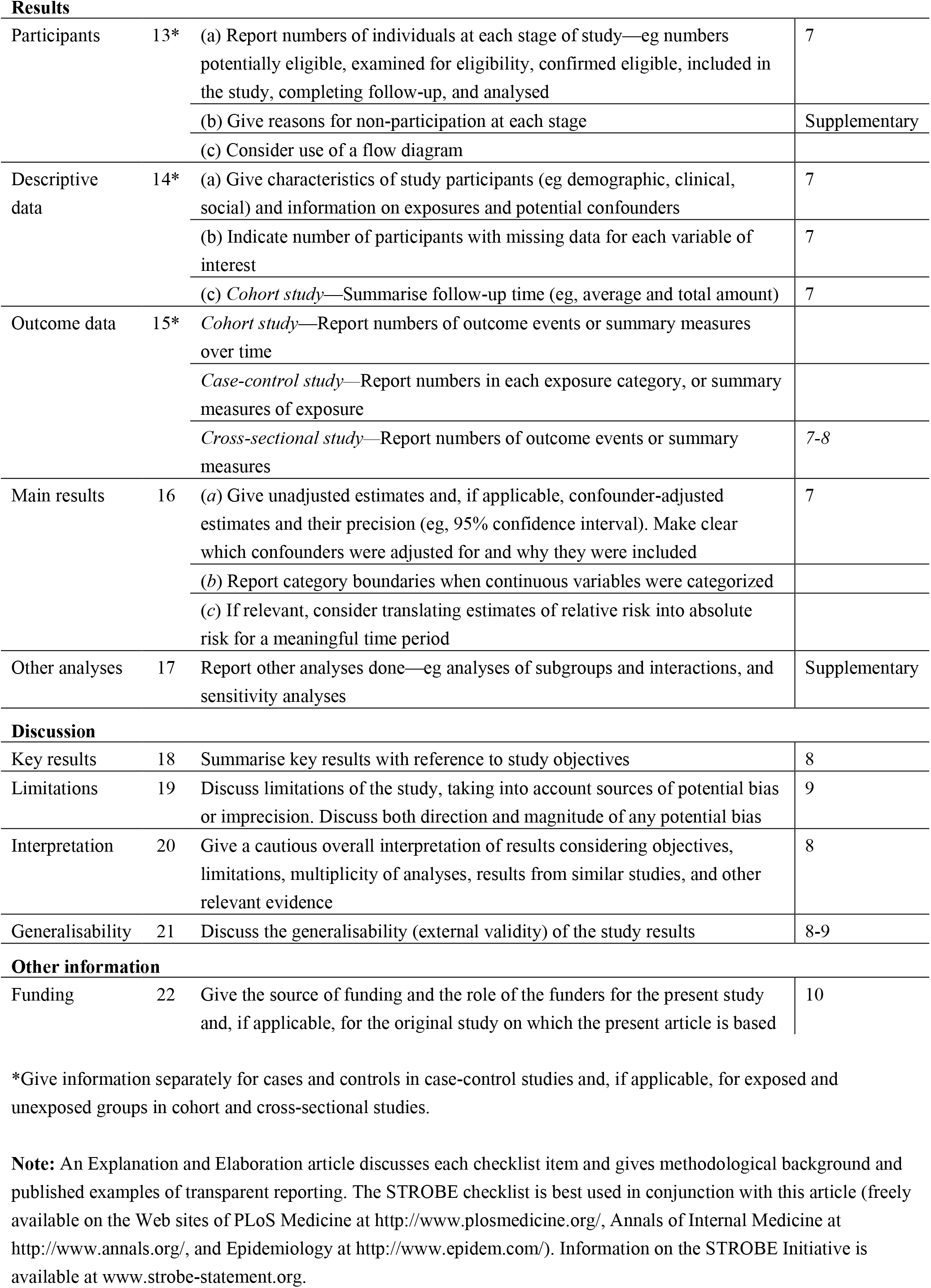

|  | Item No | Recommendation | Page No |
| --- | --- | --- | --- |
| Title and abstract | 1 | (a) Indicate the study's design with a commonly used term in the title or the abstract | 1 |
|  |  | (b) Provide in the abstract an informative and balanced summary of what was done and what was found | 2 |
| **Introduction** |  |  |  |
| Background/rationale | 2 | Explain the scientific background and rationale for the investigation being reported | 3 |
| Objectives | 3 | State specific objectives, including any prespecified hypotheses | 4 |
| **Methods** |  |  |  |
| Study design | 4 | Present key elements of study design early in the paper | 4 |
| Setting | 5 | Describe the setting, locations, and relevant dates, including periods of recruitment, exposure, follow-up, and data collection | 5 |
| Participants | 6 | (a) *Cohort study*—Give the eligibility criteria, and the sources and methods of selection of participants. Describe methods of follow-up<br>*Case-control study*—Give the eligibility criteria, and the sources and methods of case ascertainment and control selection. Give the rationale for the choice of cases and controls<br>*Cross-sectional study*—Give the eligibility criteria, and the sources and methods of selection of participants | 5 |
|  |  | (b) *Cohort study*—For matched studies, give matching criteria and number of exposed and unexposed<br>*Case-control study*—For matched studies, give matching criteria and the number of controls per case |  |
| Variables | 7 | Clearly define all outcomes, exposures, predictors, potential confounders, and effect modifiers. Give diagnostic criteria, if applicable | 6 |
| Data sources/<br>measurement | 8 | For each variable of interest, give sources of data and details of methods of assessment (measurement). Describe comparability of assessment methods if there is more than one group | 4 |
| Bias | 9 | Describe any efforts to address potential sources of bias | 9 |
| Study size | 10 | Explain how the study size was arrived at | 4-5 |
| Quantitative variables | 11 | Explain how quantitative variables were handled in the analyses. If applicable, describe which groupings were chosen and why | 6 |
| Statistical methods | 12 | (a) Describe all statistical methods, including those used to control for confounding | 6 |
|  |  | (b) Describe any methods used to examine subgroups and interactions | 4-6 |
|  |  | (c) Explain how missing data were addressed | 4-6 |
|  |  | (d) *Cohort study*—If applicable, explain how loss to follow-up was addressed<br>*Case-control study*—If applicable, explain how matching of cases and controls was addressed<br>*Cross-sectional study*—If applicable, describe analytical methods taking account of sampling strategy | 4-6 |
|  |  | (e) Describe any sensitivity analyses |  |
| **Results** |  |  |  |
| Participants | 13* | (a) Report numbers of individuals at each stage of study—eg numbers potentially eligible, examined for eligibility, confirmed eligible, included in the study, completing follow-up, and analysed | 7 |
|  |  | (b) Give reasons for non-participation at each stage | Supplementary |
|  |  | (c) Consider use of a flow diagram |  |
| Descriptive data | 14* | (a) Give characteristics of study participants (eg demographic, clinical, social) and information on exposures and potential confounders | 7 |
|  |  | (b) Indicate number of participants with missing data for each variable of interest | 7 |
|  |  | (c) *Cohort study*—Summarise follow-up time (eg, average and total amount) | 7 |
| Outcome data | 15* | *Cohort study*—Report numbers of outcome events or summary measures over time |  |
|  |  | *Case-control study*—Report numbers in each exposure category, or summary measures of exposure |  |
|  |  | *Cross-sectional study*—Report numbers of outcome events or summary measures | 7-8 |
| Main results | 16 | (a) Give unadjusted estimates and, if applicable, confounder-adjusted estimates and their precision (eg, 95% confidence interval). Make clear which confounders were adjusted for and why they were included | 7 |
|  |  | (b) Report category boundaries when continuous variables were categorized |  |
|  |  | (c) If relevant, consider translating estimates of relative risk into absolute risk for a meaningful time period |  |
| Other analyses | 17 | Report other analyses done—eg analyses of subgroups and interactions, and sensitivity analyses | Supplementary |
| **Discussion** |  |  |  |
| Key results | 18 | Summarise key results with reference to study objectives | 8 |
| Limitations | 19 | Discuss limitations of the study, taking into account sources of potential bias or imprecision. Discuss both direction and magnitude of any potential bias | 9 |
| Interpretation | 20 | Give a cautious overall interpretation of results considering objectives, limitations, multiplicity of analyses, results from similar studies, and other relevant evidence | 8 |
| Generalisability | 21 | Discuss the generalisability (external validity) of the study results | 8-9 |
| **Other information** |  |  |  |
| Funding | 22 | Give the source of funding and the role of the funders for the present study and, if applicable, for the original study on which the present article is based | 10 |

\*Give information separately for cases and controls in case-control studies and, if applicable, for exposed and unexposed groups in cohort and cross-sectional studies.
**Note:** An Explanation and Elaboration article discusses each checklist item and gives methodological background and published examples of transparent reporting. The STROBE checklist is best used in conjunction with this article (freely available on the Web sites of PLoS Medicine at http://www.plosmedicine.org/, Annals of Internal Medicine at http://www.annals.org/, and Epidemiology at http://www.epidem.com/). Information on the STROBE Initiative is available at www.strobe-statement.org.

